# Proactive vs. reactive country responses to the COVID19 pandemic shock

**DOI:** 10.1101/2021.12.06.21267351

**Authors:** Pier Luigi Sacco, Francesco Valle, Manlio De Domenico

## Abstract

The infection caused by SARS-CoV-2, responsible for the COVID-19 pandemic, is characterized by an infectious period with either asymptomatic or pre-symptomatic phases, leading to a rapid surge of mild and severe cases putting national health systems under serious stress. To avoid their collapse, and in the absence of pharmacological treatments, during the early pandemic phase countries worldwide were forced to adopt strategies, from elimination to mitigation, based on non-pharmacological interventions which, in turn, overloaded social, educational and economic systems. To date, the heterogeneity and incompleteness of data sources does not allow to quantify the multifaceted impact of the pandemic at country level and, consequently, to compare the effectiveness of country responses. Here, we tackle this challenge from a complex systems perspective, proposing a model to evaluate the impact of systemic failures in response to the pandemic shock. We use health, behavioral and economic indicators for 44 countries to build a shock index quantifying responses in terms of robustness and resilience, highlighting the crucial advantage of proactive policy and decision making styles over reactive ones.

## Introduction

In January 2020, China reported a fast increase in the number of cases of a new, severe and acute respiratory syndrome due to a novel coronavirus, the SARS-CoV-2 [1]. In less than 3 months, the pathogen spread at a global scale, leading to the COVID-19 pandemic [2, 3, 4, 5, 6]. The new disease was characterized by both asymptomatic and pre-symptomatic phases, leading to systematic under-detection of cases that threatened epidemic control [7]. The virus – exhibiting similarities with SARS and MERS pathogens [8], causing systemic disorders [9] through panviral disease mechanisms [10] – was responsible for a communicable disease characterized by a basic reproduction number of 2-3.5 [11], high enough for a sustained community transmission that could potentially overwhelm even the most prepared public health systems. In the absence of suitable medical treatments and vaccines, local governments opted for the introduction of a variety of non-pharmacological interventions (NPI), ranging from physical distancing to mandatory mask wearing, and in many cases to draconian measures such as travel bans and national lockdowns [12]. The impact of containment strategies was readily analyzed [13, 14, 11, 15, 16] to assess the relative effectiveness of available interventions [17] and identify clear change points in the time course of epidemiological signals [18].

On the one hand, some countries adopted a proactive response, devising clear anticipatory steps to take in order to achieve an elimination strategy. On the other hand, many other countries adopted a reactive response, devising procedures to avoid the overwhelming of their national health systems while allowing the virus to spread. In the following, we will refer to these two approaches as proactive vs. reactive – or, equivalently, elimination vs. mitigation – strategies, respectively. Not surprisingly, both approaches had a substantial impact on human activities and national socio-economic systems.

At the social level, the non-pharmacological measures have severely restricted social interaction with negative consequences on mental health [19], and with huge personal and collective costs [20]. At the economic level, the pandemic has caused a collapse of production and business activity, pushing many companies of any size, from very large to very small, on the brink of bankruptcy [21], and exacerbating socio-economic inequalities [22]. Such negative effects are likely to persist at least partly in the long term [23]. At the educational level, the crisis has disrupted educational programs, with especially negative impacts on students with socioeconomically fragile backgrounds [24].

### Results

The interdependency between health, social and economic systems characterizes our society as a system of systems [25, 26] which can be studied under the lens of complexity science. Any system of interest has some (unknown) structure of interdependencies that it is exposed to shocks of various nature. It is plausible to assume that perturbations will propagate through the system, altering its function – measured by some systemic indicator *S*(*t*) – through a multiplicative cascade process [27], where a change at time *t* triggers even larger changes at a subsequent time *t* + Δ*t*, resulting in a quick decrease in the value of *S*(*t*). This first phase defines the failure of a system, and it can be modeled by a logistic-like expansion (see Materials and Methods), as shown in Fig. 1A, characterized by a carrying capacity accounting for the fact that resources that can be damaged are finite. For instance, in the case of the economic system, the systemic failure can be measured by relative changes in the GDP growth and employment levels of a country. At some point, failures can no longer propagate either because there are no more resources or because mitigation procedures have been adopted: at this time step, *t*_*peak*_, the systemic indicator *S*(*t*) reaches a global minimum followed by an increase, which indicates that the system is bouncing back towards its fully operational state. This change of regime marks the beginning of a recovery phase and can be characterized by another logistic-like expansion, where one positive intervention on the system at time *t* facilitates subsequent interventions at the subsequent time *t* + Δ*t*. The overall process, detailed in Materials and Methods, leads to the model schematically reproduced in Fig. 1A, which is described by the shock function

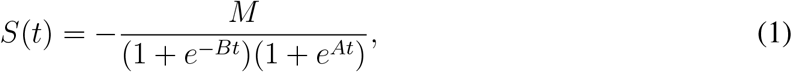

where *M, A* and *B* are parameters to be estimated from empirical observations. The area under the two curves before and after the global minimum *S*_*peak*_ = *S*(*t*_*peak*_) can be used to quantify the robustness and the resilience of the underlying system to a given shock. If we define the total damage at time *t* as

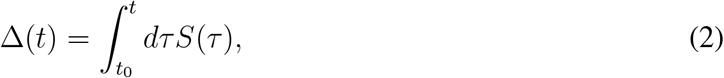

the robustness of a system is related to the share of the area covered until the peak is reached, i.e.

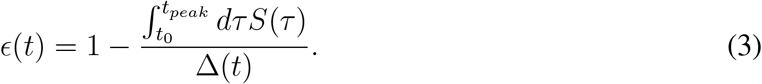

If the system had a perfect response to the shock, remaining substantially unaffected, the failure phase is characterized by an area under the curve equal to zero and the robustness index equals 1. Conversely, if the system instantaneously collapses, the robustness index equals 0. Similarly, we can define the resilience of a system by the share of the area covered after the peak is reached, i.e.

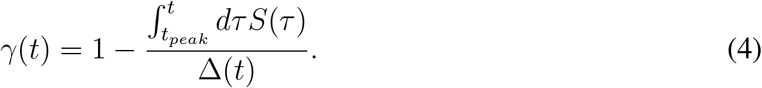

**Figure 1:**
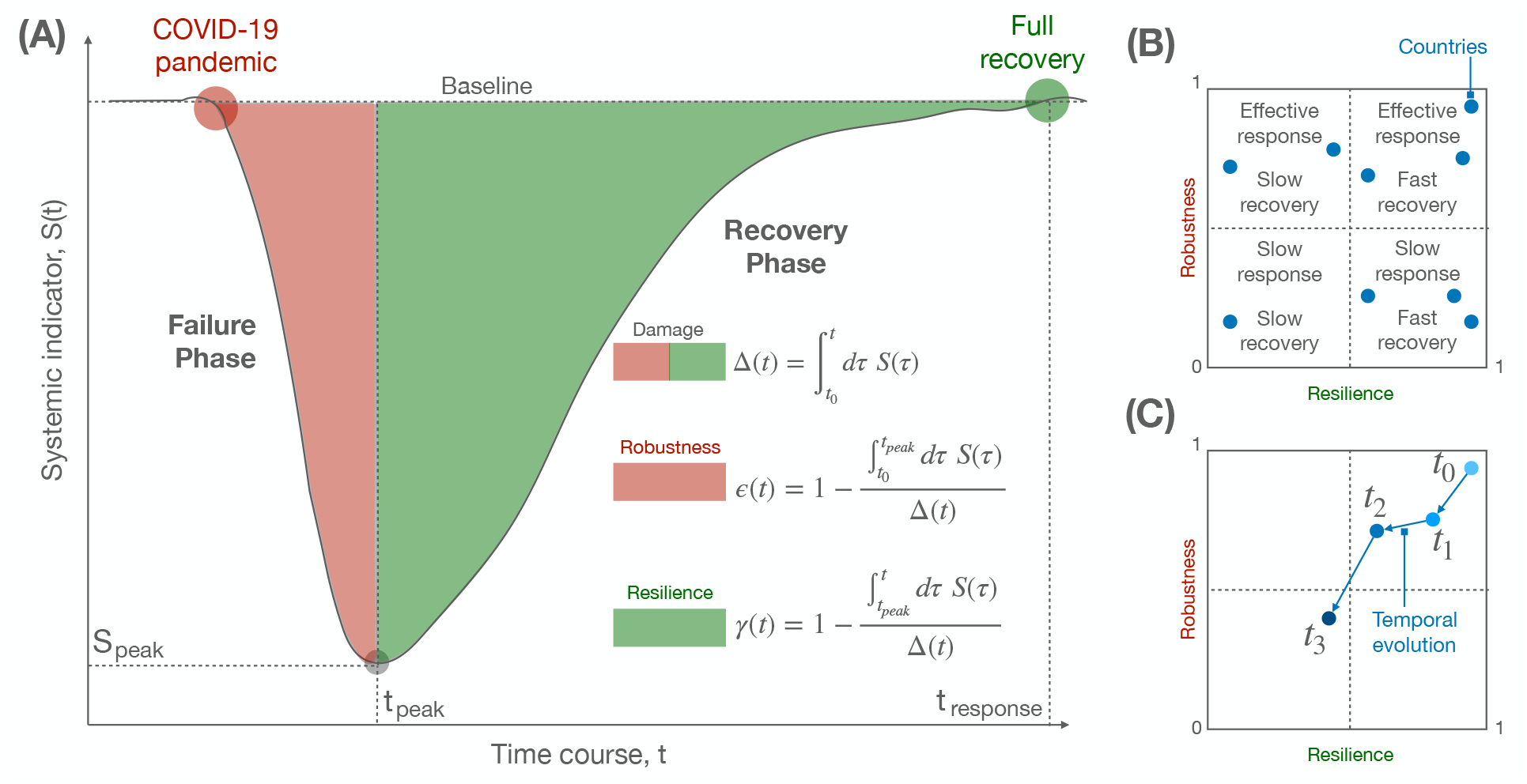
Modeling the response of a complex system to a shock. (A) Schematic of changes in a complex system – e.g., the economy of a country – as quantified by a systemic indicator – e.g., the change in GDP growth – undergoing a decrease, corresponding to a failure phase, and an increase, corresponding to a recovery phase, after a shock like COVID-19. The shaded areas under the curve allow to define a measure of robustness and resilience, which can be used to quantify the response of a country. (B) The two indices are scattered to define 4 distinct types of response within a fixed temporal window, combining effective or weak robustness with fast or slow resilience. (C) Similar to (B), but considering the temporal evolution of the two indices for a given country, allowing to monitor the trend of the system over time.

A system which instantaneously recovers to its original function will be characterized by a resilience index equal to 1. In practice, the two measures are not independent and one can expect the existence of a trade-off between robust response and fast recovery. Figure 1B shows how the relationship between robustness and resilience can be used to understand the response of different countries over a defined time window, or their evolution over time (Fig. 1C).

We consider a broad set of indicators at country level, covering health, social, behavioral and economic aspects of systemic response to the COVID-19 pandemic. Specifically, we consider closure and health indices, built by accounting for several policies such as school closing, restrictions on gatherings, testing, contract tracing, etc. (see Materials and Methods), for a total of 8 indicators for closure and 6 indicators for health as developed by the Oxford COVID-19 Government Response Tracker [28]. The behavioral index is obtained from Google human mobility anonymous and aggregate data [29] – already used to estimate optimal mobility reduction for the mitigation of COVID-19 transmission [30] – in terms of the median of the percent change from baseline value across six distinct types of movements: Retail and recreation, Grocery and pharmacy, Parks, Public transport stations, Workplaces and Residential. The economic index is built from the OECD Weekly Tracker [31], providing the percent change in weekly GDP levels from the pre-crisis trend. The epidemic index is obtained from the cumulative number of confirmed deaths due to COVID-19 relative to the total population in 2020 multiplied by one million [32].

To allow for a comparison across countries, we have normalized the observational period between February 23, 2020 and August 1, 2021, where we have information for 44 countries worldwide. As an emblematic example, we show in Fig. 2 the time course of the economic index and the composite health index for six countries, together with the corresponding shock functions, showing a nice agreement between data and expectation. However, while it is possible to compare robustness and resilience of countries with respect to a single indicator, it is still difficult to compare countries by means of a comprehensive index, to capture the multi- faceted aspects of country-level responses. To overcome this issue, for each country separately we perform a convolution of the time course of its indices to obtain an overall shock index which harmonizes the heterogeneous signals that we consider. The result of this mathematical operation is shown in Fig. 3 for two representative countries, namely Italy and New Zealand. The absolute value of the shock index in the two cases, obtained from the convolution of the aforementioned indices, reveals three orders of magnitude of difference in the overall response, with New Zealand exhibiting both higher robustness and higher resilience than Italy. We extend this analysis to all countries in our data set and show the results in Fig. 4.

**Figure 2:**
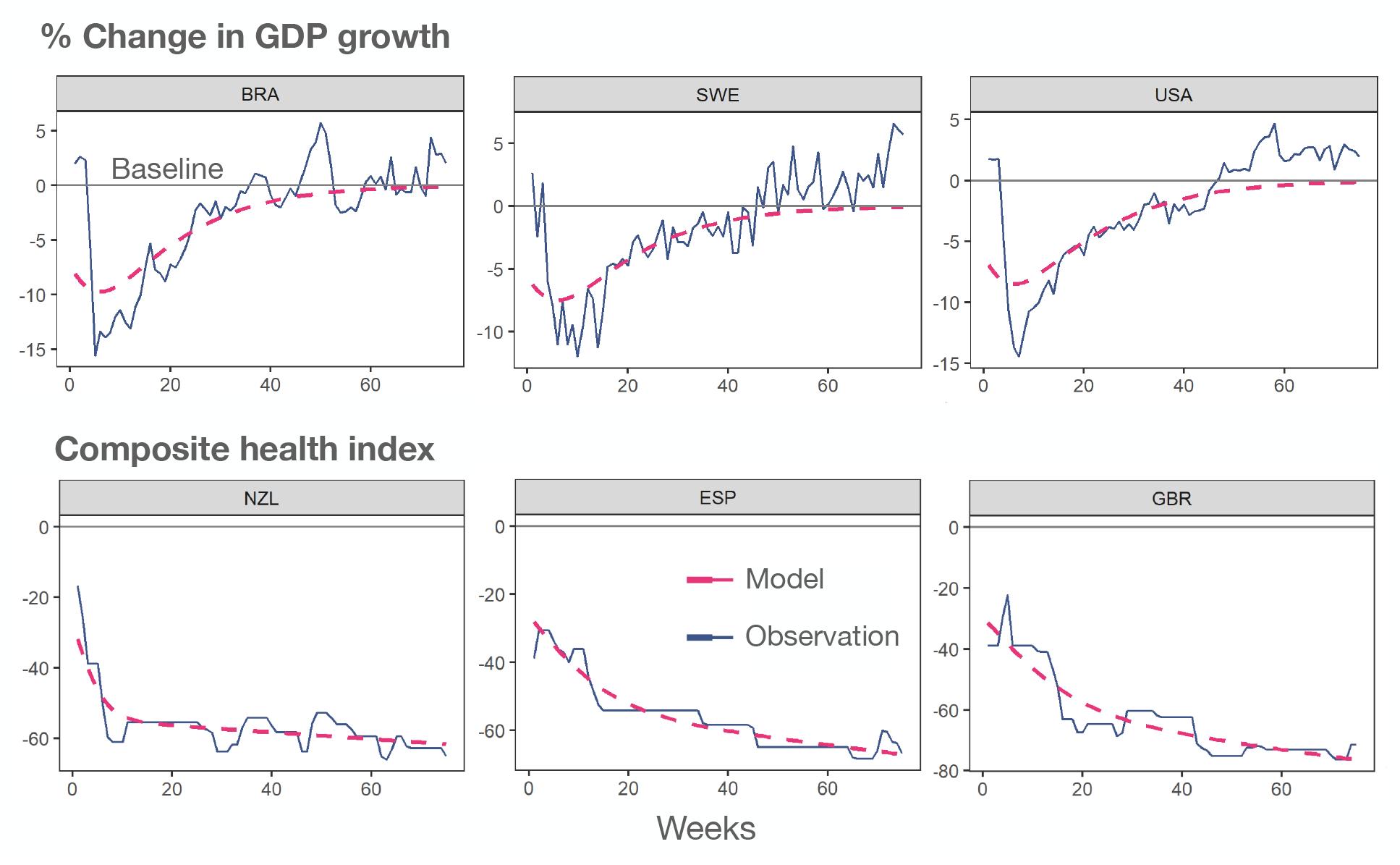
Agreement between model and country-level change indices. Observed time course of two distinct indices, economic (top) and health (bottom), for six distinct countries worldwide. The dashed line indicates the fit obtained through the corresponding shock function introduced in this work.

**Figure 3:**
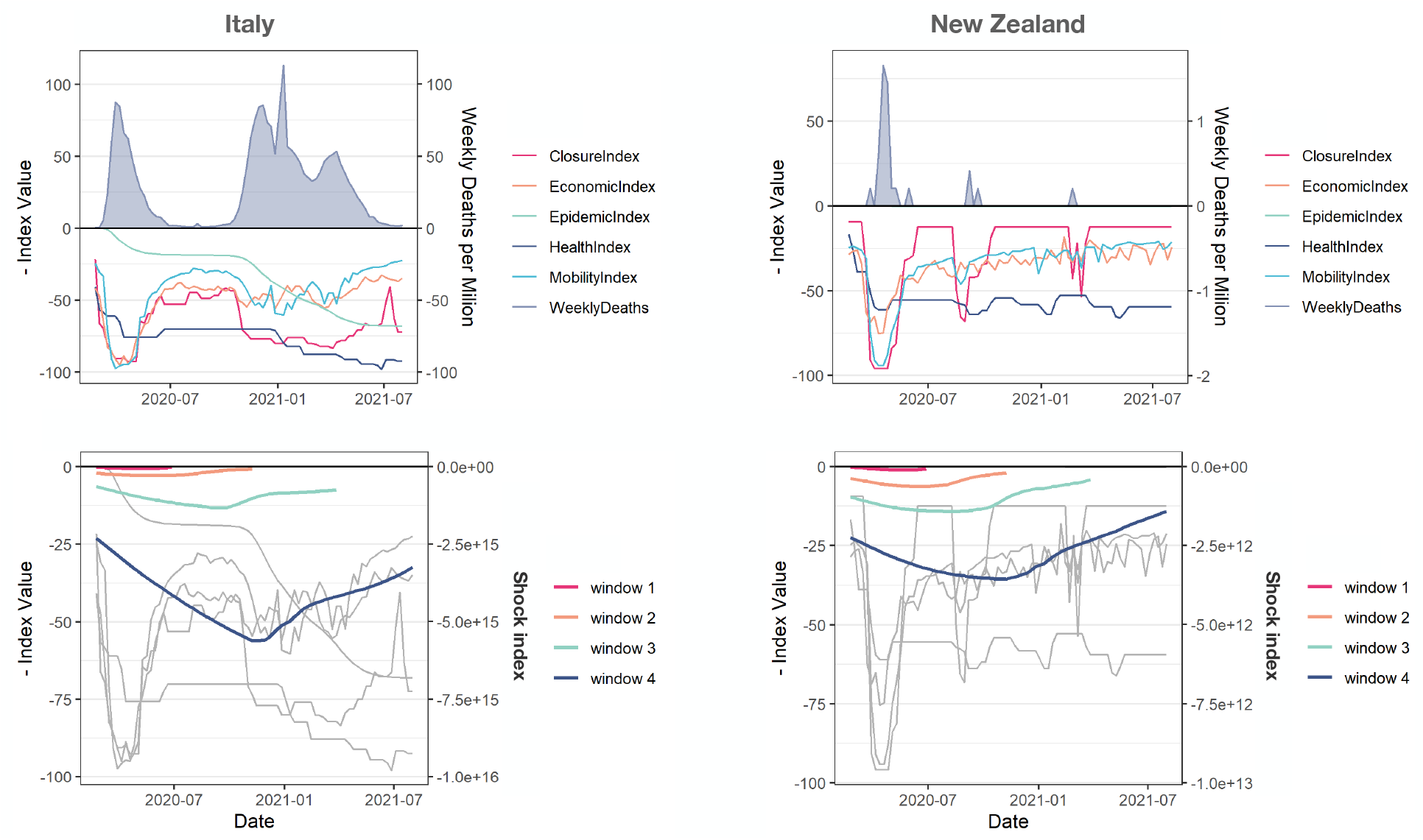
The shock index allows to compare the response of distinct countries to COVID-19. For two countries, Italy (left) and New Zealand (right), the time course of the six indices (top) and their convolution into a comprehensive shock index (bottom) is shown, for distinct values of the time window indicating a 19-weeks incremental length of the observational time between February 23, 2020 and August 01, 2021.

**Figure 4:**
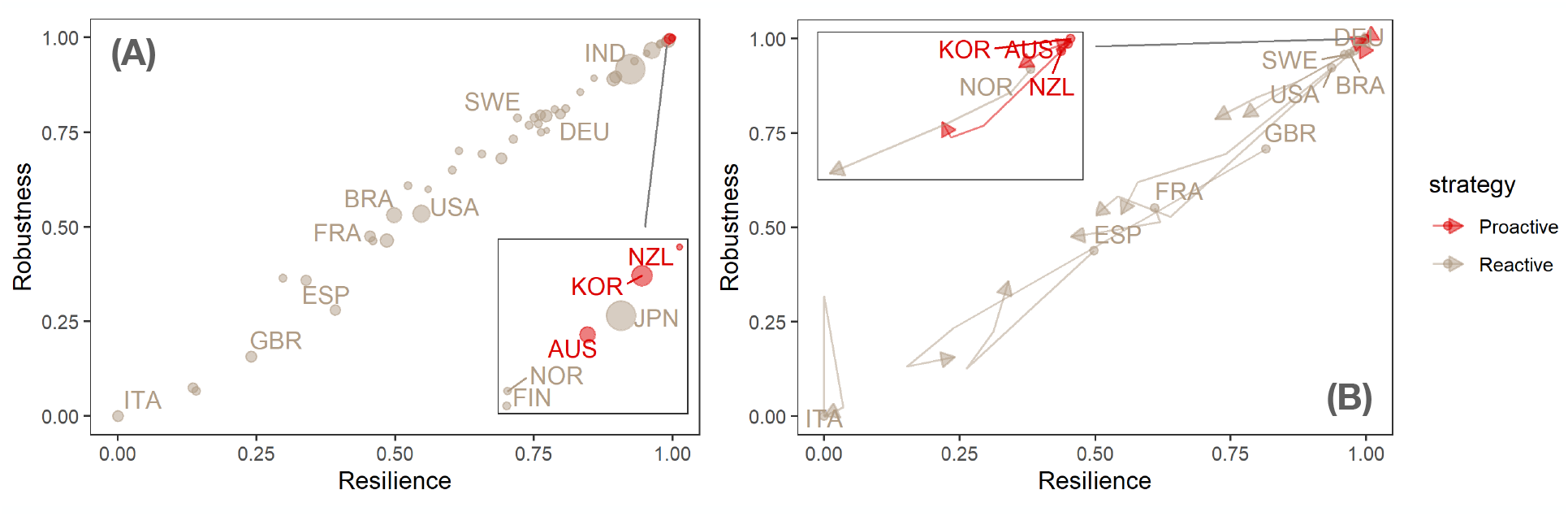
Proactive vs reactive strategies to COVID-19 pandemic. (A) static shock map where each point indicates a country and colors code proactive vs reactive response strategies. Remarkably, proactive strategies have a clear advantage, although some countries with reactive strategies, such as Nordic ones (except for Sweden) and Japan, perform similarly in robustness and resilience. (B) For a subset of countries, the evolution of their shock response over time is shown at different temporal snapshots.

Fig. 4 shows that the large majority of countries in our sample has opted for a reactive rather than a proactive strategy. However, the few countries adopting a proactive strategy consistently outperform the others both in terms of robustness and resilience. A few reactive countries feature levels of performance that are comparable to those of proactive ones. However, proactive countries better preserve their performance over time whereas similarly performing reactive ones slide down. Moreover, performances across reactive countries are very different, as shown for instance by EU countries, all of which took a reactive approach, but with very different results (Nordic countries generally do better than Mediterranean ones). The performances of proactive countries are instead very similar. Notice that geographical factors such as insularity do not have a clear effect on performance, as shown by the comparative results of the proactive New Zealand vs. the reactive UK. Likewise, performance is not critically affected by levels of socio-economic development: for instance, US and Brazil fare quite similarly, and India largely outperforms both. The fact that our model, leading to the shock function (see Materials and Methods for details), perfectly reproduces the behavior of the comprehensive shock index, provides interesting insights about the mechanisms – based on multiplicative growth processes – behind systemic failure and recovery of a country in response to external shocks such as the COVID-19 pandemic.

## Conclusions

Our results seem to deliver some clear messages that may be relevant for future policy design in response to pandemic shocks. First of all, proactive strategies seem strongly preferable to reactive ones as their immediate and anticipatory response curbs the diffusion of the virus and prevents the amplification of major socio-economic effects. Second, although in principle there should be a trade-off between robustness and resilience, our data show that in practice they are almost perfectly correlated, allowing an almost strict ranking of the performance of countries. Moreover, the performance of countries cannot be accounted for by traditional metrics such as levels of socio-economic development, and seems to depend on still poorly understood structural factors. In particular, the fact that only a few countries were able to adopt superior proactive response strategies need not depend only on political choices. Implementing a proactive strategy calls for high levels of social governance that might not be attainable in all countries without targeted adjustments. However, in view of our results, it could be advisable that countries capitalize upon the policy lessons of the current pandemic, and focus upon setting the conditions for a timely adoption of proactive responses against likely future pandemic shocks.

## Methods

Our measures of robustness and resilience are computed from a shock index built on government, health, economic and behavioral indicators combined together by iterative convolutions.

### Closure and health indices

Our government indices are based on different indicators collected by the Oxford COVID-19 Government Response Tracker team [28] and include information about restriction and containment policies as well as testing, tracking and vaccination policies. We use those indicators to create two non-overlapping indices that score between 0 and 100, exploiting the procedure described by the Oxford team, and we then apply a 7-days rolling mean on them.

### Economic index

The economic index we use is the OECD Weekly Tracker of GDP growth [31] that proxies the percent change in weekly GDP levels from the pre-crisis trend. We scaled this index between 0 and 100, where 100 is the maximum negative percent change across all countries in a given period.

### Behavioral index

The behavioral index is based on Google mobility data [29] and is computed by the median of the percent change from baseline value of retail and recreation, grocery and pharmacy, parks, public transport stations, workplaces and residential displacements. We then applied a 7-days rolling mean within each country and the resulting value is scaled between 0 and 100, where 100 is the maximum negative value across all countries in a given period.

### Epidemic index

The epidemic index for a country is equal to the cumulative number of confirmed Covid-19 deaths [28] divided by the total population in 2020 [33] multiplied by one million. Such value is then scaled between 0 and 100, where 100 is the maximum across all countries in a given period. The value of the index is retained weekly to match the granularity of the other data sources.

### Comprehensive shock index

Finally, we started by convolving two of the above indices (Closure and Economic) and then using the result as input for a second convolution with another index in an iterative fashion. Then we selected the date corresponding to the maximum value of the final convolution as a splitting point of the area under the convolution: robustness is the area under the convolution from the starting point to the “peak”, while resilience is the area under the convolution between the “peak” and the ending point of the time window considered. Both areas are estimated by numerical integration. Standardized values of robustness and resilience were then computed by subtracting from one the ratio between the value of each measure and its maximum.

Concerning the indicators dynamics, we divided our observation period in four overlapping time windows with increasing length of 19 weeks starting from February 23, 2020. For each time window we re-computed the scaled indices and the corresponding final convolution and peak date to get our robustness and resilience metrics, that were finally standardized as mentioned above.

### Mathematical model

As outlined in Fig. 1 of the main text, we characterize the evolution of a shock by means of two distinct phases:

1. *Failure phase:* where the shock induces perturbations decreasing the system’s function or, equivalently, increasing the system’s dysfunction through an expansion.
2. *Recovery phase:* where interventions to mitigate the failures induces new perturbations increasing the system’s function or, equivalently, decreasing the system’s dysfunction through a contraction.

Given the nature of complex adaptive systems, it is plausible to assume that growing and shrinking dynamics are governed by some kind of multiplicative process, which is the only fundamental assumption of our model.

During the expansion phase – where failures are detrimental for the system’s function – changes in the dysfunction indicator *𝒟*_*fail*_(*t*) are assumed to be proportional to the value of a suitably normalized measure indicator *y*(*t*), which in turn it is subject to a logistic shrink:

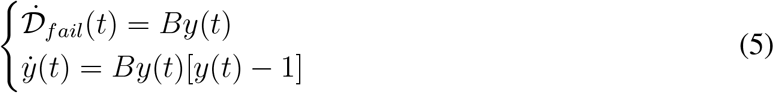

leading, for the boundary condition 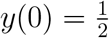, to

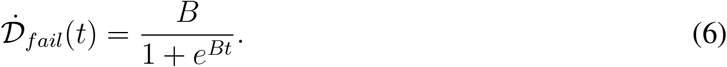

The choice of the logistic dynamics is motivated by thinking about failures in terms of a population growth which is sustained, with rate *B*, by available resources until a carrying capacity *K* is reached. To keep the model as simple as possible, we consider that *K* = 1, since maximum failures are represented by the normalized measure *y*_*max*_ = 1.

Conversely, in the contraction phase, where recovery takes place, changes in the dysfunction indicator *𝒟*_*reco*_(*t*) are assumed to be proportional to the value of a suitably normalized measure indicator *x*(*t*), which in turn it is subject to a logistic growth:

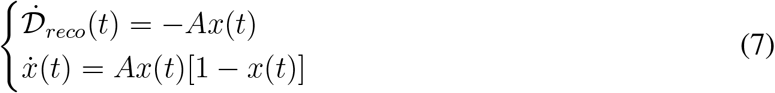

leading, for the boundary condition 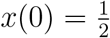, to

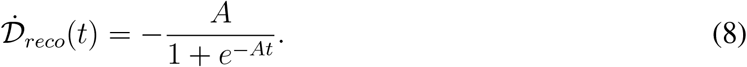

As for the failure phase, the choice of the logistic dynamics is motivated by thinking about failures in terms of a population shrink which is sustained, with rate *A*. Also in this case *K* = 1, since *x*_*max*_ = 1.

Since the two dynamics of failure and recovery compete with each other, the overall dysfunction indicator can be obtained by their additive contribution at each time *t*:

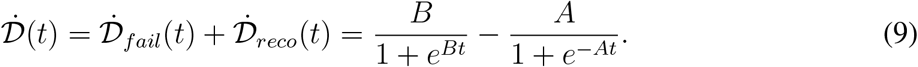

The last differential equation can be quickly solved by noting that a dysfunction indicator defined by

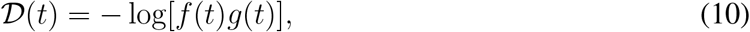

which leads to

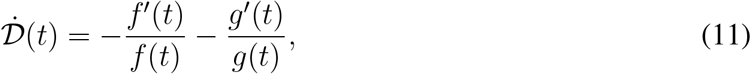

is the solution if *f* (*t*) = 1 + *e*^−*Bt*^ and *g*(*t*) = 1 + *e*^*At*^. It follows that we can write

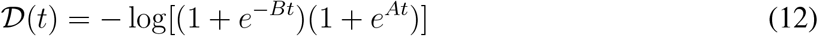

and, by introducing the **shock function** *S*(*t*) = *−Me*^*𝒟*(*t*)^, where *M* is a parameter allowing one to shift the dysfunction to account for more general deviations from the baseline, it follows

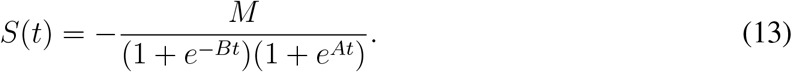

Figure 5 shows the shock function fitted to reproduce the behavior of the comprehensive shock index for a variety of countries, while Fig. 6 shows the relation between parameters *A, B* and *M*. Table 1 reports the values of the parameters for all countries analyzed in this study.

**Figure 5:**
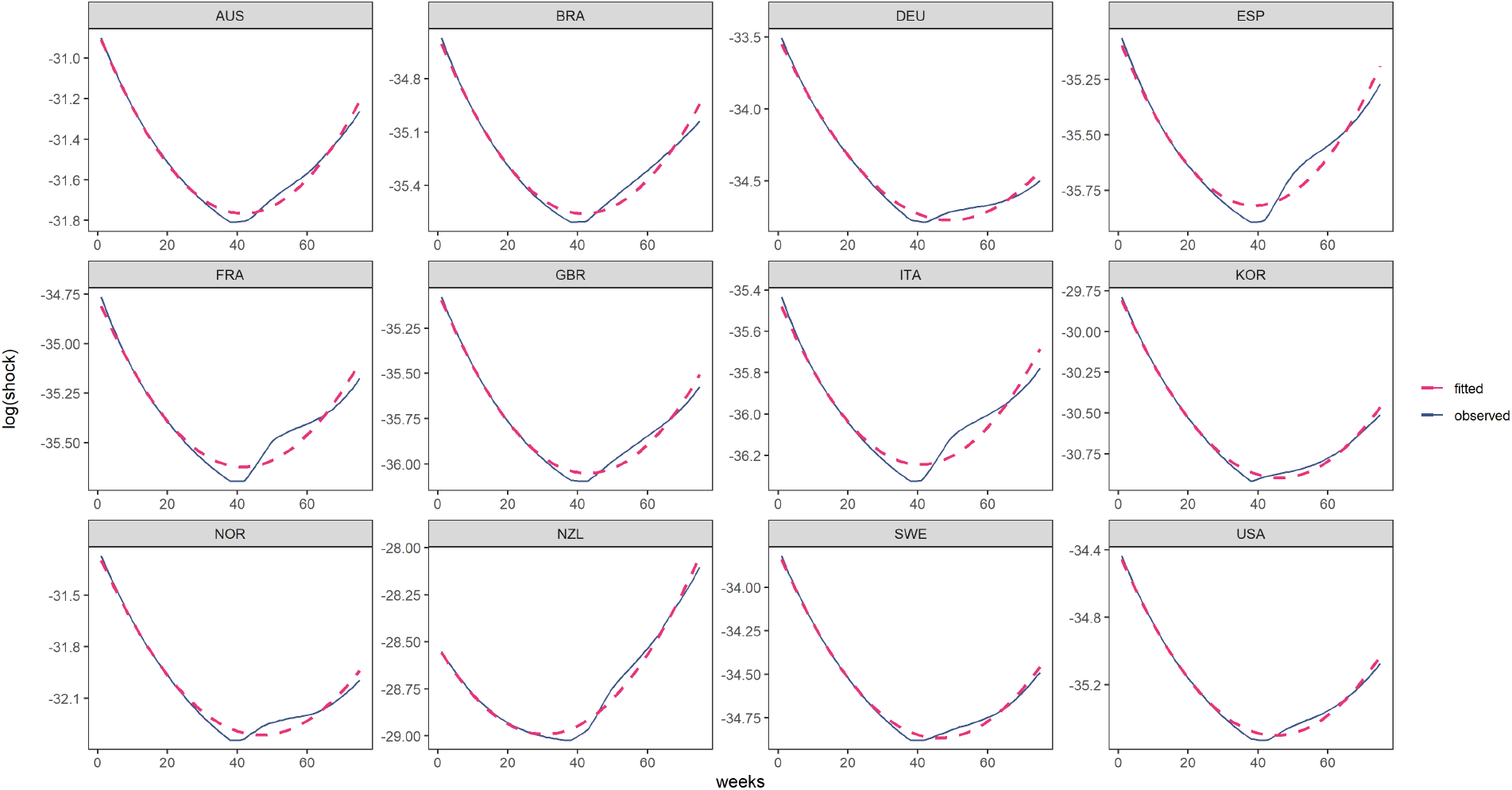
Observed time course of the global shock index for different countries. Dashed lines represent the corresponding shock function fit obtained by non-linear least squares fitting method. On the y-axis the natural logarithm of the shock index is used.

**Figure 6:**
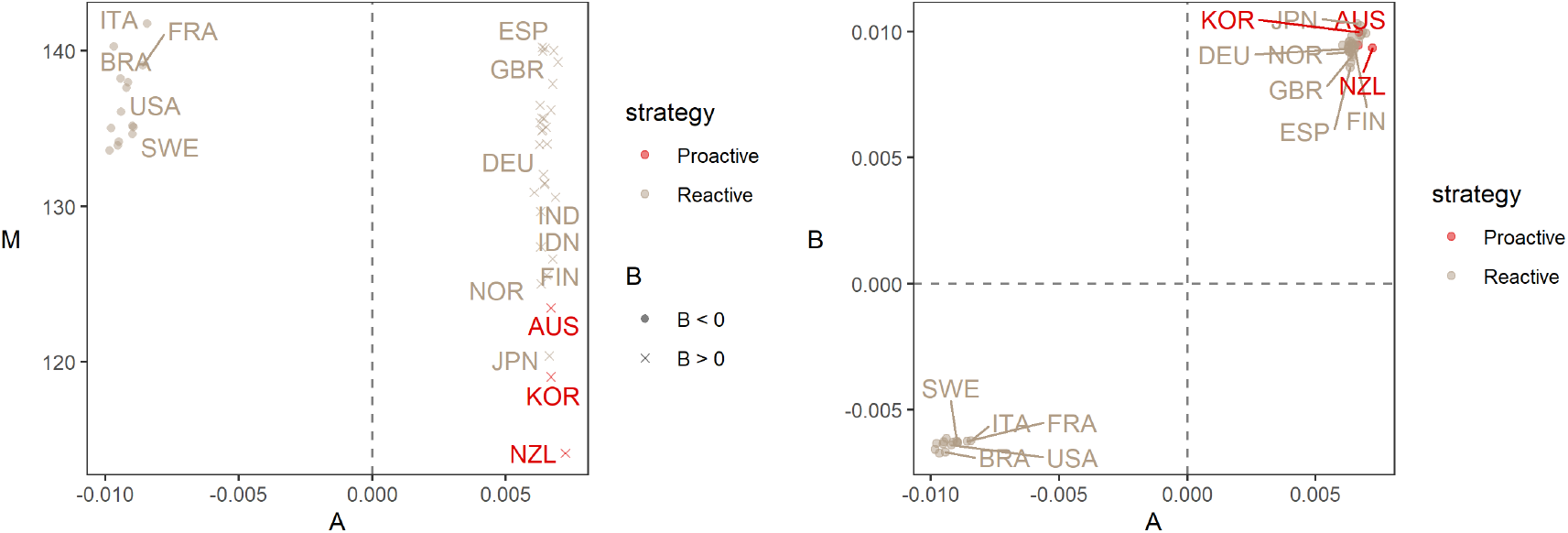
Model parameters. Estimates of model parameters for each country, obtained by non-linear least squares analysis on the log transformation of the global shock index. Gray dashed lines indicate values equal to zero. Left: scatter plot of *A* vs *B*. Right: scatter plot of *A* vs *M*, with different shapes for the sign of the parameter *B*. Colours distinguish *Proactive* from *Reactive* strategies.

**Table 1:** Model parameters. Numerical estimates of model parameters for each country, obtained by non-linear least squares analysis on the log transformation of the global shock index. BIC indicates the Bayesian Information Criterion indicator.

| Country | M | B | A | BIC | Strategy |
| --- | --- | --- | --- | --- | --- |
| ARG | 140.03 | 0.0099 | 0.0068 | -296.85 | Reactive |
| AUS | 123.47 | 0.0095 | 0.0067 | -326.54 | Proactive |
| AUT | 135.39 | 0.0094 | 0.0063 | -238.84 | Reactive |
| BEL | 139.97 | 0.0088 | 0.0064 | -277.35 | Reactive |
| BGR | 133.61 | -0.0066 | -0.0098 | -199.95 | Reactive |
| BRA | 138.22 | -0.0067 | -0.0094 | -269.45 | Reactive |
| CAN | 134.64 | -0.0063 | -0.009 | -285.32 | Reactive |
| CHE | 134.86 | 0.0092 | 0.0063 | -282.64 | Reactive |
| CHL | 140.29 | -0.0067 | -0.0097 | -334.4 | Reactive |
| COL | 139.3 | 0.0099 | 0.007 | -286.43 | Reactive |
| CZE | 136.1 | -0.0061 | -0.0094 | -233.93 | Reactive |
| DEU | 134 | 0.0093 | 0.0063 | -269.15 | Reactive |
| DNK | 129.67 | 0.0092 | 0.0063 | -290.78 | Reactive |
| ESP | 140.23 | 0.0086 | 0.0064 | -232.99 | Reactive |
| EST | 127.4 | 0.0096 | 0.0063 | -212.74 | Reactive |
| FIN | 125.57 | 0.0093 | 0.0065 | -286.16 | Reactive |
| FRA | 139.08 | -0.0063 | -0.0086 | -221.81 | Reactive |
| GBR | 140.2 | 0.0091 | 0.0065 | -293.18 | Reactive |
| GRC | 134.15 | -0.0063 | -0.0095 | -186.44 | Reactive |
| HUN | 135.71 | 0.0096 | 0.0064 | -215.38 | Reactive |
| IDN | 126.62 | 0.0102 | 0.0068 | -250.72 | Reactive |
| IND | 130.58 | 0.01 | 0.0069 | -260.91 | Reactive |
| IRL | 135.66 | 0.009 | 0.0064 | -275.85 | Reactive |
| ISR | 134.02 | 0.0095 | 0.0065 | -275.63 | Reactive |
| ITA | 141.77 | -0.0062 | -0.0084 | -216.55 | Reactive |
| JPN | 120.39 | 0.0103 | 0.0066 | -287.05 | Reactive |
| KOR | 119.04 | 0.01 | 0.0067 | -346.1 | Proactive |
| LTU | 132.06 | 0.0098 | 0.0064 | -221.29 | Reactive |
| LUX | 135.1 | 0.0093 | 0.0065 | -247.36 | Reactive |
| LVA | 130.9 | 0.0094 | 0.0061 | -261.43 | Reactive |
| MEX | 137.9 | 0.0098 | 0.0068 | -323.22 | Reactive |
| NLD | 135.11 | -0.0063 | -0.0089 | -313.11 | Reactive |
| NOR | 125.01 | 0.0092 | 0.0063 | -270.93 | Reactive |
| NZL | 114.11 | 0.0094 | 0.0072 | -289.59 | Proactive |
| POL | 133.92 | -0.0064 | -0.0095 | -204.01 | Reactive |
| PRT | 137.98 | -0.0063 | -0.0091 | -212.78 | Reactive |
| ROU | 134.93 | 0.0095 | 0.0064 | -210.98 | Reactive |
| RUS | 131.45 | 0.0095 | 0.0064 | -261.13 | Reactive |
| SVK | 135.04 | -0.0063 | -0.0098 | -211.91 | Reactive |
| SVN | 136.51 | 0.0094 | 0.0063 | -239.4 | Reactive |
| SWE | 135.17 | -0.0063 | -0.009 | -342.45 | Reactive |
| TUR | 131.49 | 0.0094 | 0.0065 | -221.61 | Reactive |
| USA | 137.63 | -0.0064 | -0.0092 | -327.78 | Reactive |
| ZAF | 136.21 | 0.0096 | 0.0067 | -266.25 | Reactive |

## Data Availability

All data produced in the present work are contained in the manuscript and cited publicly available repositories.

https://www.google.com/covid19/mobility/

https://ourworldindata.org/coronavirus

https://github.com/OxCGRT/covid-policy-tracker

